# Olfactory impairment and the risk of major adverse cardiovascular outcomes in older adults

**DOI:** 10.1101/2023.10.27.23297697

**Authors:** Keran W. Chamberlin, Yaqun Yuan, Chenxi Li, Zhehui Luo, Mathew Reeves, Anna Kucharska-Newton, Jayant M. Pinto, Jiantao Ma, Eleanor M. Simonsick, Honglei Chen

**Author notes:** Corresponding author: Department of Epidemiology and Biostatistics, College of Human Medicine, Michigan State University, East Lansing, MI.

## Abstract

**Background:** Olfactory impairment is common in older adults and may be associated with adverse cardiovascular health; however, empirical evidence is sparse.

**Objective:** To examine olfaction and the risk of coronary heart disease (CHD), stroke, and congestive heart failure (CHF).

**Methods:** This study included 2,537 older adults (aged 75.6±2.8 years) from the Health ABC Study with olfaction assessed by the 12-item Brief Smell Identification Test in 1999-2000, defined as poor (score ≤8), moderate (9-10), or good (11-12). The outcomes were incident CHD, stroke, and CHF.

**Results:** During up to 12-year follow-up, 353 incident CHD, 258 stroke, and 477 CHF events were identified. Olfaction was associated with incident CHF, but not with CHD or stroke. After adjusting for demographics, the cause-specific hazard ratio (HR) of CHF was 1.35 (95% confidence interval (CI): 1.08, 1.70) for moderate and 1.39 (95%CI: 1.09, 1.76) for poor olfaction. With additional adjustment for lifestyle, chronic diseases, and biomarkers of CHF, the HR was modestly attenuated to 1.32 (95%CI: 1.05, 1.66) for moderate and 1.28 (95%CI: 1.01, 1.64) for poor olfaction. These associations were robust in pre-planned subgroup analyses by age, sex, race, and prevalent CHD/stroke. However, the associations appeared to be evident among participants who reported very-good-to-excellent health (HR=1.47 (95%CI: 1.02, 2.13) for moderate and 1.76, (95%CI: 1.20, 2.57) for poor olfaction). In contrast, null association with CHF was found among those with fair-to-poor self-reported health.

**Conclusions:** In community-dwelling older adults, a single olfaction test was associated with a long-term risk for incident CHF, particularly among those reporting very-good-to-excellent health.

## Background

The human sense of smell declines with age in older adults. Prevalence of poor olfaction, assessed by smell-identification screening, quickly increases from ∼6% in age 50s to over 60% in 80s^1^. This age-dependent olfactory decline has been confirmed in multiple community-based studies^2,3^. Despite the high prevalence of poor olfaction in older adults, our understanding of its health implications has been largely limited to its role as a prodromal symptom of neurodegeneration and its robust association with mortality^4^. Interestingly, our recent findings^5^ indicated that only 22% of the excess mortality associated with poor olfaction could be explained by dementia and Parkinson’s disease (PD), suggesting that poor olfaction may have more profound health implications than what is known to date. Further, this association with higher mortality was limited to participants who self-reported good-to-excellent health at baseline^5^, raising the possibility that poor olfaction may be a marker of deteriorating health that precedes the emergence of more traditionally recognized signs and symptoms of health decline.

Beyond neurodegenerative diseases and mortality, the health implications of poor olfaction have been subject to wide speculation, with limited empirical evidence. Recent data suggest that poor olfaction is associated with carotid intima-media thickness and artery plaques^6,7^, suggesting that smell loss may be a marker of atherosclerosis – the underlying pathogenesis of cardiovascular disease. Further, poor olfaction may gradually degrade one’s food choices, adversely affecting dietary quality and nutrition^8,9^, which may contribute to cardiovascular disease over time. Therefore, as a nonspecific subclinical marker and/or a potential contributor, poor olfaction may be related to cardiovascular risk. Because poor olfaction is prevalent among older adults and cardiovascular disease is the leading cause of death and disability, this potential association should be investigated further.

To the best of our knowledge, only one study^10^ has prospectively examined the association between olfaction and heart diseases, and reported an elevated but statistically non-significant association with heart attack and/or heart disease. We hereby comprehensively examined olfaction status in relation to the risk of three major adverse cardiovascular conditions − coronary heart disease (CHD), stroke, and congestive heart failure (CHF) among community-dwelling older adult participants in the Health, Aging, and Body Composition (Health ABC) Study.

## Methods

### Study population

The Health ABC Study aims to investigate the interrelationships among aging-related conditions, social and behavioral factors, and physiological and functional changes in older adults^11^. Briefly, in 1997 and 1998, this study recruited 3,075 well-functioning, community-dwelling older adults (51.5% women and 41.7% blacks) aged 70-79 years in the designated zip code areas surrounding Pittsburgh, Pennsylvania, and Memphis, Tennessee. Eligibility criteria included no reported difficulty in walking 1/3 mile or climbing up 10 steps, no active fatal cancers, and no plans to move in 3 years. Study participants were followed with annual clinic visits through Year 6, and then in Year 8, 10, 11, and 16. Phone interviews were conducted to update health status every 6 months until Year 15 and then quarterly through Year 17. In the current analysis, we used the Year-3 clinic visit (1999-2000) as the baseline which was when the olfaction test was conducted. The primary analysis was limited to 2,537 participants after excluding those who missed Year-3 clinic visit (n=154) and did not take the smell test (n=384).

In the analysis of each cardiovascular outcome, we excluded prevalent cases of each outcome at baseline, respectively. As case adjudications for major health outcomes (except death) were conducted through August 14, 2012, we followed at-risk participants from baseline until the first cardiovascular outcome, death, last contact, or the end of the 12-year follow-up, whichever came first. The Health ABC study protocol was approved by all relevant institutional review boards, and all participants provided written informed consent at enrollment.

### The Brief-Smell Identification Test (B-SIT)

Olfaction was tested at the Year-3 clinic visit, using the 12-item cross-cultural B-SIT. This test is a shortened version of the 40-item University of Pennsylvania Smell Identification Test and has been widely used in large populations^12^. The 12-item test is brief, convenient, and well-suited to field settings in large epidemiological studies and quick clinical screening^13,14^. Participants were instructed to smell each of the 12 odorants, one at a time, and then to identify the odorant from 4 possible answers in a forced multiple-choice format. One point was given for each correct answer with a total score ranging from 0 to 12. We defined poor olfaction as a B-SIT score ≤8, moderate as 9-10, and good as 11-12, approximately corresponding to the tertile distribution of the B-SIT testing score in the study population. Using these cut-points, we have reported strong associations of poor olfaction with PD, dementia, total mortality, and pneumonia hospitalization in this cohort.

### Major adverse cardiovascular outcomes

The Health ABC study closely monitored the health and survival of study participants via study clinic visits, semiannual phone updates, and surveillance of hospitalization and death. As detailed previously ^15–18^, major adverse cardiovascular outcomes were first identified via the cohort’s routine follow-ups and health surveillance and then adjudicated according to a standard protocol. Briefly, at each clinic visit and semi-annual telephone interview, participants or their proxies were asked directed questions about cardiovascular disease events diagnosed by a physician, overnight hospitalizations, and outpatient cardiovascular procedures such as angioplasty since the last interview. Once an event was reported, local medical event adjudicators collected and reviewed related medical records according to a standardized study protocol^19^. For each death event, study investigators had an exit interview with a knowledgeable proxy who provided detailed information on the death event and the participant’s physical functioning before death. The immediate and underlying causes were adjudicated centrally by an expert committee after reviewing hospital records, death certificates, autopsy findings, and informant interviews.

In this study, we defined incident CHD as the first event of myocardial infarction (MI), angina pectoris, or death with CHD as the underlying cause. According to the protocol^19^, MI adjudication accounted for evolving diagnostic electrocardiogram (ECG) pattern, diagnostic ECG pattern, and abnormal cardiac enzymes, or ischemic symptoms and either an evolving ST-T pattern or an obscure ECG pattern. The adjudication of angina pectoris considered symptoms such as chest pain, chest tightness, shortness of breath, and a diagnosis from a physician, as well as medical treatment including nitroglycerin, beta-blocker, or calcium channel blocker. We defined stroke as the first event of stroke or death with cerebrovascular diseases as the underlying cause, considering evidence of a rapid onset of neurologic deficit attributed to obstruction or rupture of the arterial system and new CT/MRI lesion consistent with clinical presentation of stroke without evidence of alternative causes (e.g., tumor or infection). We defined CHF as the first admission of overnight hospitalization with CHF adjudicated as the primary inpatient reason or a concurrent event. The adjudication considered physician diagnosis, and medical treatments for CHF including both a diuretic and digitalis or a vasodilator, or the presence of cardiomegaly and pulmonary edema on chest X-ray, or evidence of a dilated ventricle and global/ segmental wall motion abnormalities with deceased systolic function either by ECG or contrast ventriculography.

### Covariates

As few risk factors have been established for olfactory loss in older adults except for age, sex, and race, we mainly considered cardiovascular risk factors/predictors as covariates in the analyses. The adjustment of these covariates may help control for potential confounding and improve statistical efficiency^20^. With few exceptions, we used covariate data from the Year-3 clinic visit when the smell testing was conducted. Age, sex, race, study site, education level, smoking status, minutes of brisk walking per week, and general health status were self-reported. Body mass index (BMI) was calculated by dividing weight by height-squared (kg/m^2^) and systolic blood pressure by averaging two measures in the sitting position. The use of antihypertensive medication was assessed using the medication inventory method coded with the Iowa Drug Information System Drug Vocabulary and Thesaurus^21^. We defined comorbidities according to published protocols, in brief, 1) diabetes as self-reported diagnosis by a physician, the use of anti-diabetic drugs, a fasting blood glucose level of ≥126 mg/dL, or an oral glucose tolerance test of ≥200mg/dL^22^; 2) dementia as the score of the Modified Mini-Mental State examination (3MS) at the Year-1 clinic visit less than 80, a decline in 3MS score from Year 1 through Year 3 at least 1.5 race-stratified standard deviations, an adjudicated diagnosis of dementia based on hospitalization, or documented medication uses for dementia ^5^; 3) PD as adjudicated by two movement disorder specialists by consensus after review of self-reported diagnosis by a physician, medication uses, hospitalization records, and adjudicated cause of death^23^. Depressive symptoms were defined as a score of ≥10 on the Center for Epidemiologic Studies Depression Scale Short form^24^. When covariate data are not available for the Year-3 clinic visit, we used data from previous years. Resting heart rate was measured at Year 1. Left ventricular hypertrophy (LVH) was diagnosed using Year-1 ECG according to the Minnesota code criteria^16^. Abnormal lung function was defined as the forced expiratory volume in the 1^st^ second measured at Year 1 below the lower limit of normal according to the age-, sex-and race-specific normalized reference values of the National Health and Nutrition Examination Survey Ⅲ equations^25^. Plasma total cholesterol (TC) and high-density lipoprotein-cholesterol (HDL-C) were measured using fasting EDTA plasma collected at Year 2 and Year 1, respectively^26^. Serum albumin was measured using samples collected at Year 1^27^, interleukin 6 using samples collected at Year 2^28^, and cystatin C and creatinine using samples collected at Year 3^29^. All these biomarkers have been widely analyzed in the Health ABC Study with details reported previously. We estimated the glomerular filtration rate (eGFR) mainly using the Chronic Kidney Disease Epidemiology Collaboration (CKD-EPI) creatinine-cystatin C equation; for 9.6% of the sample with missing creatinine data, we estimated eGFR using the CKD-EPI cystatin C equation^30^. Among those with creatinine measures, these two eGFR estimates were highly correlated with a Spearman coefficient of 0.82.

### Statistical analysis

In descriptive analyses, we used linear regression for continuous variables and logistic or multinomial regressions for categorical variables to estimate their age-adjusted marginal means/percentages in each olfaction group. We then calculated the cumulative incidence function (CIF) of each type of cardiovascular outcome and its corresponding competing risk of death, and tested the equality of CIF across baseline olfaction status using the Gray’s test. In multivariable analyses, we used the Cox cause-specific hazard model to account for the competing risk of death and reported cause-specific hazard ratio (HR) and 95% confidence interval (CI) for each type of cardiovascular event. This approach quantifies the direct association between olfaction status and each outcome of interest, not affected by the association of olfaction with death, fitting our analytical goal^31^. In the analyses, we first controlled for age, sex, race, education, and study site (model 1), and then further adjusted for key lifestyle cardiovascular risk factors in model 2^32,33^, including smoking status, brisk walking, BMI, self-reported general health status, antihypertensive medication use, diabetes, depressive symptoms, systolic blood pressure, TC, and HDL-C. As prevalent atherosclerotic diseases could be an important risk factor for CHF^34^, we also included prevalent CHD and stroke in model 2 of the CHF analysis. Finally, we constructed model 3 for CHF by further adjusting for previously identified markers of CHF in the cohort^16–18^, including LVH, abnormal lung function, heart rate, serum albumin, interleukin 6, and eGFR. In all regression analyses, we applied the Supremum Test to check the proportional hazard assumption and, when applicable, stratified the covariates that did not satisfy the assumption in the regression model. Finally, given the strong association of olfaction with dementia and PD, we conducted a sensitivity analysis by excluding participants with prevalent dementia or PD at baseline.

For outcomes that showed a significant association with olfaction in the primary analysis, we conducted secondary subgroup analyses by age, sex, race, self-reported health status, and history of other major cardiovascular diseases at baseline. These analyses were pre-planned because the prevalence of poor olfaction is age-dependent and substantially higher in men than in women and in blacks than in whites, and our prior analysis showed that the association of poor olfaction with higher mortality was limited to people with self-reported good-to-excellent health at baseline^5^. Interestingly, for CHF, we found that the association was evident mostly among individuals who self-reported very-good-to-excellent health. We therefore conducted two post hoc exploratory analyses in this subgroup. First, we reestimated the full model in this subgroup. Next, we further modeled the B-SIT score on a continuous scale, the non-linearity form of which was regressed by using the quadratic term. We used the SAS software (version 9.4; SAS Institute, Cary, NC) for all the analyses with a two-sided α of 0.05.

## Results

At baseline, participants were, on average, 75.6±2.8 years old, with 51.6% female and 38.5% Black. In the overall study sample, compared with participants with good olfaction, those with poor olfaction were more likely to be older, men, Black, smokers, and from Memphis (**Table 1**). They were also more likely to report a high-school education level or less and fair-to-poor general health status, and to have diabetes, abnormal or missing lung function, lower BMI, TC, HDL-C, and eGFR. As age is the most important risk factor for olfactory loss in older adults, we also presented age-adjusted covariates by olfaction in Supplementary **Table S1.** Once age was adjusted, the imbalances of prevalent diabetes, lung function, BMI, and cholesterol level across olfaction groups disappeared. Finally, we also presented population characteristics separately for each outcome of interest with the corresponding prevalent cases removed (Supplementary **Table S2-S4**), and the data were generally comparable to those in **Table 1**.

**Table 1.** Population characteristics by baseline olfaction status (n=2,537) ^a^.

| Variable | Olfaction status |  |  |
| --- | --- | --- | --- |
|  | Good<br>(n = 845) | Moderate<br>(n = 867) | Poor<br>(n = 825) |
| Age in years, median (IQR) | 75.0 (4.0) | 75.0 (5.0) | 76.0 (4.0) |
| <75 years | 391 (46.3) | 370 (42.7) | 279 (33.8) |
| ≥75 years | 454 (53.7) | 497 (57.3) | 546 (66.2) |
| Male sex, n (%) | 324 (38.3) | 418 (48.2) | 485 (58.8) |
| Black race, n (%) | 265 (31.4) | 331 (38.2) | 380 (46.1) |
| Study site, n (%) |  |  |  |
| Memphis | 377 (44.6) | 432 (49.8) | 428 (51.9) |
| Pittsburgh | 468 (55.4) | 435 (50.2) | 397 (48.1) |
| Education, n (%) |  |  |  |
| ≤high school | 414 (49.0) | 490 (56.5) | 510 (61.8) |
| >high school | 431 (51.0) | 377 (43.5) | 315 (38.2) |
| Body mass index, n (%) |  |  |  |
| <25 kg/m<sup>2</sup> | 273 (32.3) | 270 (31.1) | 313 (37.9) |
| 25-30 kg/m<sup>2</sup> | 366 (43.3) | 363 (41.9) | 338 (41.0) |
| >30 kg/m<sup>2</sup> | 206 (24.4) | 234 (27.0) | 174 (21.1) |
| Smoking status, n (%) |  |  |  |
| Never | 429 (50.8) | 390 (45.0) | 351 (42.5) |
| Former & <30 pack-years | 235 (27.8) | 217 (25.0) | 215 (26.1) |
| Current or ≥30 pack-years | 181 (21.4) | 260 (30.0) | 259 (31.4) |
| Brisk walking, n (%) |  |  |  |
| <90 min/wk | 744 (88.0) | 786 (90.7) | 754 (91.4) |
| ≥90 min/wk | 101 (12.0) | 81 (9.3) | 71 (8.6) |
| General health status, n (%) |  |  |  |
| Very good to excellent | 423 (50.1) | 384 (44.3) | 325 (39.4) |
| Good | 292 (34.6) | 351 (40.5) | 312 (37.8) |
| Fair to poor | 130 (15.4) | 132 (15.2) | 188 (22.8) |
| Systolic blood pressure in mmHg, median (IQR) | 134 (26) | 134 (24) | 134 (28) |
| Antihypertensive drug use, n (%) | 497 (58.8) | 533 (61.5) | 483 (58.5) |
| Diabetes, n (%) | 181 (21.4) | 214 (24.7) | 220 (26.7) |
| Depressive symptoms, n (%) | 86 (10.2) | 108 (12.5) | 115 (13.9) |
| Heart rate in beats per minute, median (IQR) | 63 (14) | 63 (14) | 65 (16) |
| LVH, n (%) | 98 (11.6) | 97 (11.2) | 96 (11.6) |
| Abnormal lung function, n (%) |  |  |  |
| No | 689 (81.5) | 682 (78.7) | 610 (73.9) |
| Yes | 75 (8.9) | 104 (12.0) | 104 (12.6) |
| Missing | 81 (9.6) | 81 (9.3) | 111 (13.5) |
| Total cholesterol in mg/dL, median (IQR) | 206.0 (51.0) | 204.0 (49.0) | 202.0 (51.0) |
| HDL-C in mg/dL, median (IQR) | 52.0 (19.0) | 51.0 (21.0) | 51.0 (20.0) |
| Albumin in g/dL, median (IQR) | 4.0 (0.4) | 4.0 (0.4) | 4.0 (0.5) |
| Interleukin 6 in pg/mL, median (IQR) | 2.27 (2.15) | 2.33 (2.49) | 2.33 (2.24) |
| eGFR in mL/min/1.73m<sup>2</sup>, median (IQR) | 81.1 (24.3) | 81.3 (24.5) | 77.3 (26.6) |
| Prevalent major cardiovascular diseases |  |  |  |
| Prevalent CHD, n (%) | 199 (23.6) | 207 (23.9) | 207 (25.1) |
| Prevalent stroke, n (%) | 69 (8.2) | 73 (8.4) | 65 (7.9) |
| Prevalent CHF, n (%) | 37 (4.4) | 44 (5.1) | 35 (4.2) |

Abbreviations: IQR: inter-quartile range; HDL-C: high-density lipoprotein-cholesterol; LVH: left ventricular hypertrophy; eGFR: estimated glomerular filtration rate; CHD: coronary heart diseases; CHF: congestive heart failure.
<sup>a</sup> Prevalent cases of outcomes of interest are included in this table. Please see Supplementary Materials for tables for each outcome of interest without corresponding prevalent cases.

After excluding prevalent cases at baseline, 1,924 participants were at risk for incident CHD, 2,330 for stroke, and 2,421 for CHF. During 12 years of follow-up, 353 individuals (18.3%) had an incident CHD event, 258 (11.1%) experienced an incident stroke, and 477 (19.7%) had an incident CHF hospitalization. In the descriptive analysis (**Figure 1**), baseline olfaction status was not associated with the cumulative incidence of CHD or stroke. In contrast, compared to participants with good olfaction, those with moderate and poor olfaction had a higher cumulative incidence of CHF. In all analyses, poor olfaction was associated with a higher competing risk of death.

**Figure 1.**
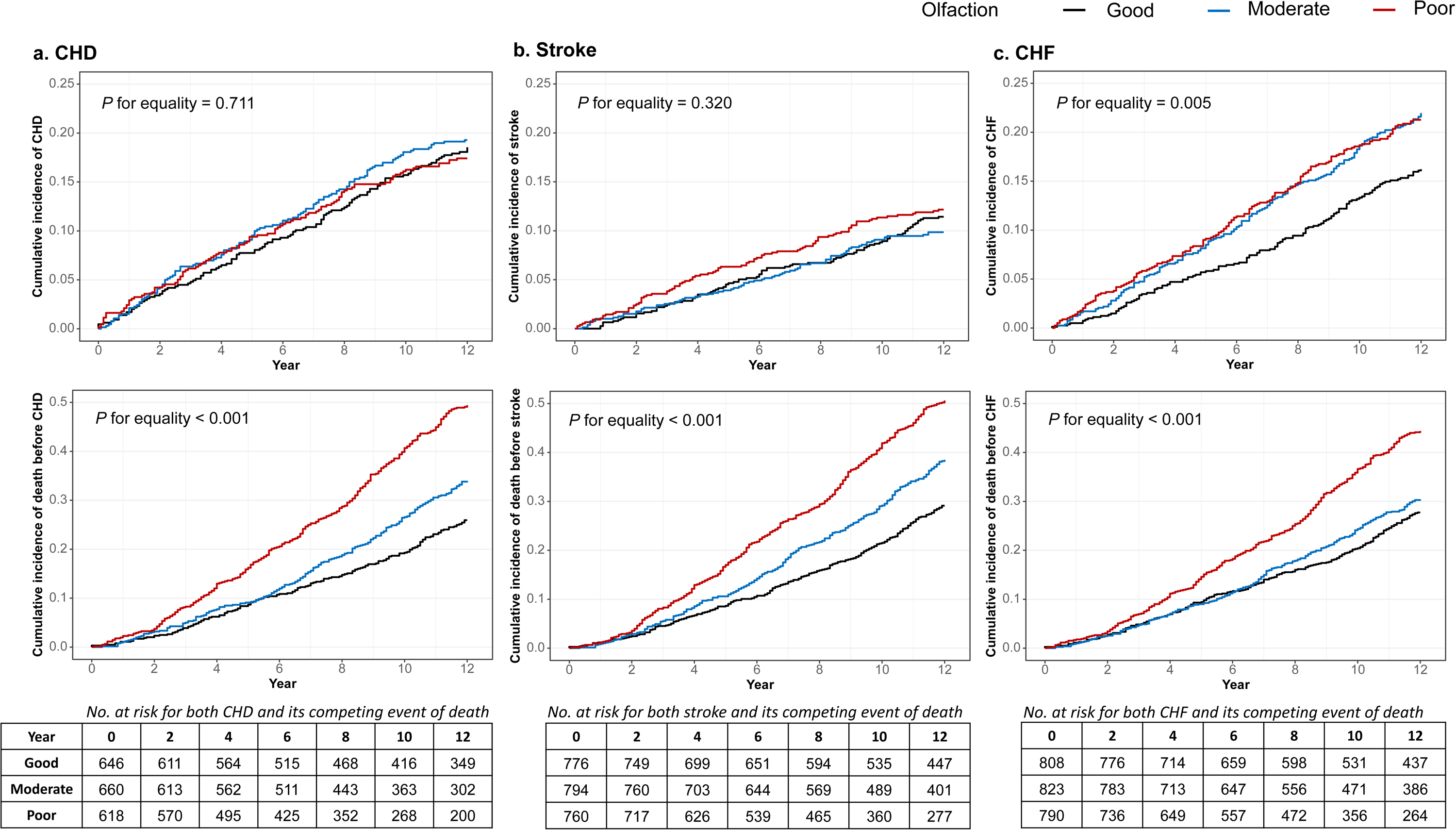
Cumulative incidence function by baseline olfaction status (good, moderate, poor) of a) coronary heart diseases (CHD) and its competing event of death (n=1,924); b) stroke and its competing event of death (n=2,330); c) congestive heart failure (CHF) and its competing event of death (n=2,421)

Multivariable models confirmed the unadjusted findings (**Table 2**). After adjusting for demographics, compared to participants with good olfaction, the cause-specific HR of CHF during a median 10.8 years of follow-up was 1.35 (95% CI: 1.08,1.70) for those with moderate olfaction and 1.39 (95% CI: 1.09, 1.76) for those with poor olfaction. The associations were barely changed with further adjustment for lifestyle risk factors and prevalent CHD/stroke, and were only modestly attenuated after further adjusting for ECG-based, spirometry-based, and blood-based biomarkers for CHF. In the fully adjusted model, the HR became 1.32 (95% CI: 1.05, 1.66) for moderate vs. good olfaction and 1.28 (95%CI: 1.01, 1.64) for poor vs. good olfaction. As in the descriptive analyses, neither CHD nor stroke outcome was associated with baseline olfaction status. After removing prevalent cases of dementia or PD at baseline, the results were consistent with our primary findings (Supplementary **Table S5**).

**Table 2.** The association of baseline olfaction status with incident coronary heart diseases (CHD), stroke, and congestive heart failure (CHF) for up to 12 years of follow-up ^a^.

| Olfactory function | No. of Event | Person-years | Incidence (per 1,000 person-year) | Model 1 <sup>b</sup> |  | Model 2 <sup>c</sup> |  | Model 3 <sup>d</sup> |  |
| --- | --- | --- | --- | --- | --- | --- | --- | --- | --- |
|  |  |  |  | HR (95% CI) | *P* | HR (95% CI) | *P* | HR (95% CI) | *P* |
| CHD (n=1,924) |  |  |  |  |  |  |  |  |  |
| Good | 119 | 6124 | 19.4 | Reference |  | Reference |  |  |  |
| Moderate | 127 | 5939 | 21.4 | 1.06 (0.83,1.37) | 0.635 | 1.01 (0.78,1.31) | 0.936 |  |  |
| Poor | 107 | 5018 | 21.3 | 1.01 (0.77,1.33) | 0.921 | 0.97 (0.73,1.27) | 0.813 |  |  |
| Stroke (n=2,330) |  |  |  |  |  |  |  |  |  |
| Good | 88 | 7665 | 11.5 | Reference |  | Reference <sup>e</sup> |  |  |  |
| Moderate | 78 | 7514 | 10.4 | 0.85 (0.62,1.15) | 0.290 | 0.85 (0.62,1.16) | 0.297 |  |  |
| Poor | 92 | 6428 | 14.3 | 1.10 (0.81,1.50) | 0.529 | 1.12 (0.82,1.52) | 0.475 |  |  |
| CHF (n=2,421) |  |  |  |  |  |  |  |  |  |
| Good | 130 | 7792 | 16.7 | Reference |  | Reference |  | Reference <sup>f</sup> |  |
| Moderate | 180 | 7533 | 23.9 | 1.35 (1.08,1.70) | 0.009 | 1.31 (1.05,1.65) | 0.020 | 1.32 (1.05,1.66) | 0.017 |
| Poor | 167 | 6561 | 25.5 | 1.39 (1.09,1.76) | 0.007 | 1.37 (1.08,1.74) | 0.010 | 1.28 (1.01,1.64) | 0.042 |

Abbreviations: HR: hazard ratio; 95% CI: 95% confidence interval
<sup>a</sup> Associations were estimated from Cox cause-specific models to account for the competing risk of death.
<sup>b</sup> Model 1 included age, sex, race, education, and study site as covariates.
<sup>c</sup> Model 2 further included smoking status, brisk walking, body mass index, self-reported general health status, systolic blood pressure, use of antihypertensive medications, diabetes, depressive symptoms, total cholesterol, and high-density lipoprotein-cholesterol as covariates. For CHF, Model 2 further included prevalent CHD/stroke in addition to the above covariates.
<sup>d</sup> Model 3 (only for CHF) further included heart rate, left ventricular hypertrophy, abnormal lung function, albumin, interleukin 6, and estimated glomerular filtration rate.
<sup>e</sup> Brisk walking and antihypertensive medication use were stratified in the Cox model.
<sup>f</sup> Tertile of interleukin 6 was stratified in the Cox model.

The associations of olfaction with CHF were robust across subgroups of age, sex, race, and baseline history of CHD and stroke (**Figure 2**). Although we did not observe a statistically significant interaction, the association of poor olfaction and CHF appears to be more evident and monotonic among participants with very-good-to-excellent health at baseline. In contrast, we found a null association among participants who self-reported fair-to-poor health.

**Figure 2.**
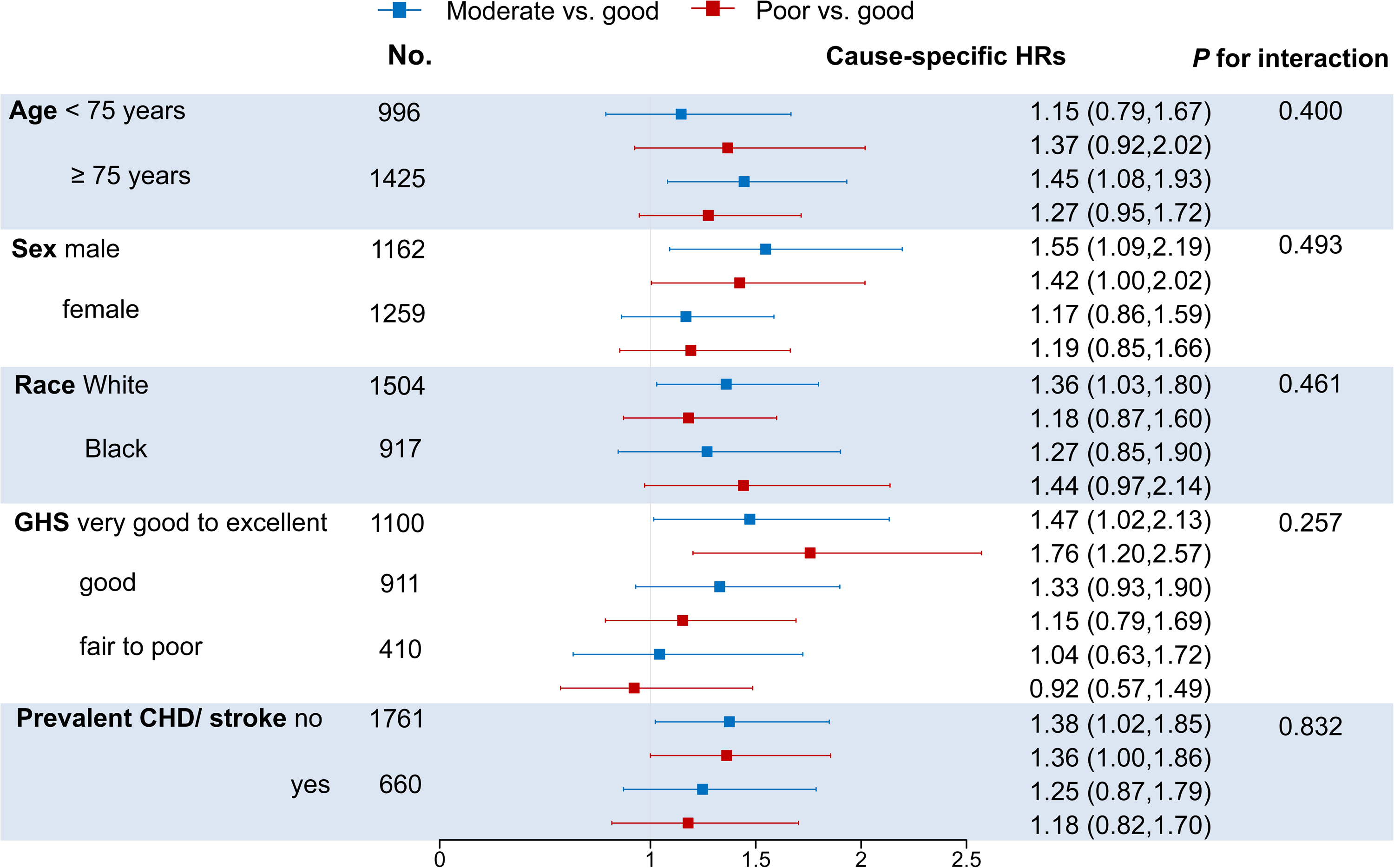
Cause-specific hazard ratios (HRs) and 95% confidence intervals (CIs) of olfaction in relation to congestive heart failure with up to 12 years of follow-up in subgroup analyses (n=2,421). Each model was adjusted for the interaction between baseline olfaction status and the subgroup factor of interest, plus covariates of age, sex, race, education, study site, smoking status, brisk walking, body mass index, self-reported general health status, systolic blood pressure, use of antihypertensive medications, diabetes, depressive symptoms, total cholesterol, high-density lipoprotein-cholesterol, prevalent coronary heart diseases (CHD)/stroke, heart rate, left ventricular hypertrophy, abnormal lung function, albumin and estimated glomerular filtration rate, stratified by the tertile of interleukin 6

We, therefore, further explored details of this relationship among participants who self-reported a very-good-to-excellent health at baseline. When we analyzed the B-SIT score as a continuous variable using the perfect score of 12 as the reference, the cause-specific HR of CHF ascended as the olfaction performance decreased until the B-SIT score of 4, after which the HRs were slightly attenuated (**Figure 3**).

**Figure 3.**
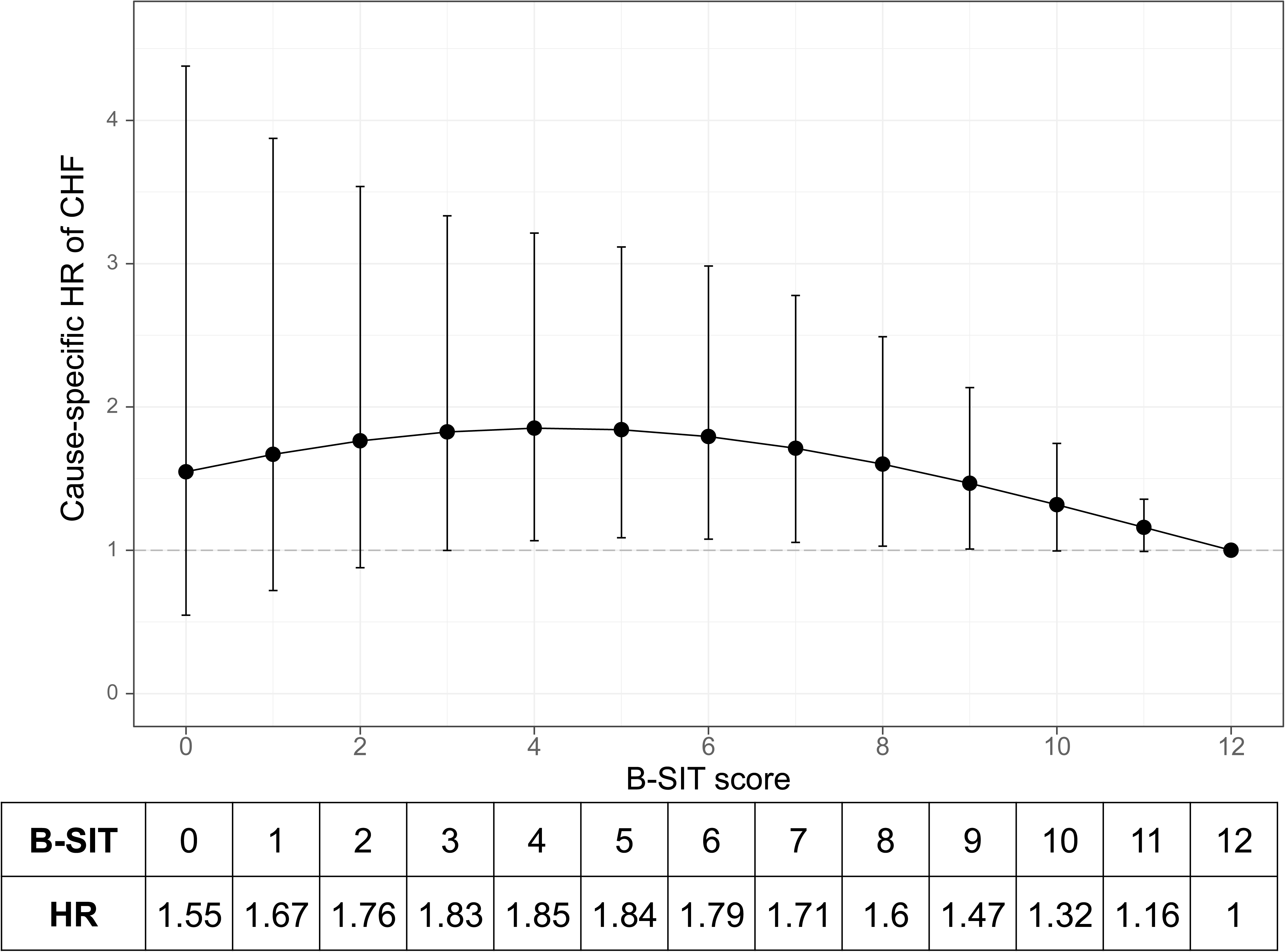
Cause-specific hazard ratios (HRs) and 95% confidence intervals (CIs) for congestive heart failure (CHF) by continuous olfaction score among participants who self-reported very-good-to-excellent health (n=1,100). Olfaction was measured by the Brief-Smell Identification test (B-SIT), the perfect score of which as 12 was used to be the reference. The model was adjusted for age, sex, race, education, study site, smoking status, brisk walking, body mass index, use of antihypertensive medications, diabetes, depressive symptoms, total cholesterol, high-density lipoprotein-cholesterol, prevalent coronary heart disease/stroke, heart rate, left ventricular hypertrophy, abnormal lung function, albumin, interleukin 6 and estimated glomerular filtration rate, stratified by groups of systolic blood pressure (140mmHg as the cut-off)

## Discussion

To our knowledge, this is the first study that aims to examine the association of olfaction with major cardiovascular diseases among older adults. Such an investigation is important because poor olfaction is prevalent in older adults, cardiovascular disease is the leading cause of death, and their connections are biologically plausible. In this large community-dwelling cohort, we found that a single test of olfaction status was associated with the risk of developing CHF for up to 12 years of follow-up. This association was robust across subgroups of age, sex, race, and prevalent CHD/stroke, but appeared to be more evident among participants who reported very-good-to-excellent health at baseline. In contrast, olfaction status was not associated with the risk of developing CHD or stroke. Taken together, this study provides interesting preliminary evidence that poor olfaction may be associated with long-term CHF risk in older adults, particularly among those who consider their general health as very good or excellent.

Cardiovascular diseases are the leading causes of death worldwide and its risk factors are among the best characterized^35^. However, as people age, known associations with cardiovascular diseases may attenuate, possibly due to aging and resilience to existing risk factors among survivors^36^. There remains a critical need to identify novel factors associated with adverse cardiovascular outcomes in older adults to further inform risk prediction and intervention. In contrast to known cardiovascular risk factors, including hypertension, obesity, and smoking, even health-conscious individuals rarely pay attention to their sense of smell^1,3^. However, poor olfaction affects about a quarter of older adults with health implications likely beyond its known associations with neurodegenerative diseases^5^.

In older adults, poor olfaction may be related to cardiovascular health either as a subclinical marker or a potential risk factor. As vascular remodeling develops and progresses, insufficient blood supply may gradually impair the health of nasal epithelium and structures in the olfactory signal pathway, limiting normal olfactory functioning^37^. Supporting this viewpoint, preliminary evidence suggests that carotid intima-media thickness and artery plaques, two subclinical markers of atherosclerosis, have been associated with the olfactory decline in older adults^6,7^. On the other hand, it is also biologically plausible that poor olfaction may adversely affect cardiovascular health. It has been speculated that impaired olfaction may alter ones’ diet and food choices, which could negatively impact their nutritional status and overall health over time^8,9^. Further, impaired olfaction may contribute to a depressed mood, social isolation, and physical inactivity^38^. All these may potentially increase one’s vulnerability to endogenous and exogenous stressors, contributing to or exacerbating cardiovascular disease risk. However, to date, the role of olfaction in cardiovascular health remains speculative with limited direct empirical evidence.

To the best of our knowledge, only one prospective study has explored the association of poor olfaction with the risk of cardiovascular diseases. In the National Social Life, Health, and Aging Project, Siegel et al reported that olfactory decline during the first 5 years was associated with marginally significantly greater odds of reporting a diagnosis of heart disease (odds ratio: 1.75, 95% CI: 0.93–3.31)^10^. Notably, in this study, the diagnosis of heart disease was self-reported only once at the year-10 follow-up survey and was analyzed as a secondary outcome. The current study is large, community-based, and specifically designed to examine olfaction in relation to the risk of adjudicated incident CHD, stroke, and CHF. In the analyses, we carefully accounted for the competing risk of death and relevant covariates. We found that a single smell test was not associated with incident CHD or stroke events. However, compared to participants with good olfaction, those with moderate or poor olfaction had a robust, albeit modest, increase in CHF risk for up to 12 years of follow-up. In contrast to CHD and ischemic stroke where arterio-and/or atherosclerosis are major mechanisms^39,40^, CHF is etiologically more complex. The latter is a multiorgan syndrome with a net outcome of a failing heart, characterized by a reduced cardiac output and increased venous pressure^41^. Coronary arteriosclerosis contributes to CHF, but any sustained myocardial stress such as increased cardiac pressure and volume overload may lead to myocardial hypertrophic response and cardiac remodeling, eventually resulting in CHF^34^. Although speculative, the differential results of CHF from CHD/stroke support the possibility that poor olfaction may signal or elevate one’s vulnerability to myocardial stressors.

Interestingly, the association of poor olfaction with incident CHF appeared to be more evident among participants who self-reported very-good-to-excellent health status at baseline, similar to our finding on the association of poor olfaction with mortality^5^. In contrast, we found a null association among individuals who self-evaluated their overall health as fair or poor. Self-reported health is a subjective perception that one may integrate their biological, social, mental, and functional health perspectives with their personal and cultural beliefs and their attitudes towards health^42^. While the report is subject to individual interpretation, it has been commonly used in health research to assess one’s general health status and it robustly predicts the risk of mortality in older adults^43^. Although our analysis is mostly exploratory, we speculated individuals who rated their health as fair or poor might already have multiple comorbidities or risk factors that play a detrimental role in their myocardial health, outweighing that from poor olfaction. In contrast, among those who reported very-good-to-excellent health, poor olfaction may serve as an early signal for deteriorating myocardial health in the absence of other clinical signals for increased CHF risk. Notably, in this subgroup, the association estimate of poor olfaction with CHF was modestly higher than that of known leading causes of CHF such as prevalent cardiovascular disease and habitual smoking (Supplementary **Table S6**). Taken together, we speculate poor olfaction is likely an early indicator for deteriorating myocardial health in apparently healthy older adults, awaiting independent confirmation and investigation of underlying mechanisms.

Strengths of this study include the relatively large number of community-based participants, more than a decade of follow-up, meticulous health surveillance and outcome assessments, careful covariate identification, and statistical analyses. Our study also has several notable limitations. First, study participants were all older than 70 at enrollment but were relatively high-functioning. Therefore, study findings may not be readily generalizable to younger populations or populations with different demographics or health status. Second, olfaction was only assessed once. As olfactory function declines fast with age in older adults, participants’ olfaction may continuously decline over follow-up, which was not captured in the current study. We therefore might have underestimated the role of olfaction in signifying or maintaining cardiovascular health in older adults. Future longitudinal studies with relatively younger participants and repeated assessments of olfaction may better characterize the role of olfaction in cardiovascular health in the context of aging. Third, as the B-SIT was designed to screen for smell identification deficit in large populations, our study did not address the association of other olfactory modalities (e.g., detection and discrimination) with the risk of major cardiovascular outcomes. Fourth, while our study findings were robust as evidenced in multiple sensitivity analyses, as in any observational study, we could not exclude the possibilities of chance finding or residual confounding. Finally, while our study suggests that both poor and moderate olfaction are associated with the future risk of experiencing a CHF event, it provides little clue to the underlying mechanisms.

In conclusion, in this well-established community-based study of older adults, we found that poor olfaction was associated with risk for CHF for up to twelve years, but not with risk for CHD or stroke. Future studies should confirm this observation and investigate underlying mechanisms.

## Acknowledgments

We thank the participants of the Health ABC study for their contributions and dedication to health research.

## Author’s contributions

HC and KWC conceived the study. HC and EMS provided the data. KWC, CL, ZL, and HC designed the study and codnucted statistical analyses. KWC and HC prepared the first draft of the manuscript and all other coauthors provided critical revision comments. All authors accessed and verified data. All authors contributed to the interpretation of the results and approved the final version for submission. HC had final responsibility to submit for publication.

## Data Availability Statement

Data used in this study are available from the National Institute on Aging (NIA) Health ABC Study. To access the data, investigators should submit an analytical proposal online at https://healthabc.nia.nih.gov/ancillary-biospecimen-proposals.

